# Using next generation matrices to estimate the proportion of infections that are not detected in an outbreak

**DOI:** 10.1101/2021.02.24.21252339

**Authors:** H Juliette T Unwin, Anne Cori, Natsuko Imai, Katy A. M. Gaythorpe, Sangeeta Bhatia, Lorenzo Cattarino, Christl A. Donnelly, Neil M. Ferguson, Marc Baguelin

## Abstract

Contact tracing, where exposed individuals are followed up to break ongoing transmission chains, is a key pillar of outbreak response for infectious disease outbreaks. Unfortunately, these systems are not fully effective, and infections can still go undetected as people may not remember all their contacts or contacts may not be traced successfully. A large proportion of undetected infections suggests poor contact tracing and surveillance systems, which could be a potential area of improvement for a disease response. In this paper, we present a method for estimating the proportion of infections that are not detected during an outbreak. Our method uses next generation matrices that are parameterized by linked contact tracing data and case line-lists. We validate the method using simulated data from an individual-based model and then investigate two case studies: the proportion of undetected infections in the SARS-CoV-2 outbreak in New Zealand during 2020 and the Ebola epidemic in Guinea during 2014. We estimate that only 5.26% of SARS-CoV-2 infections were not detected in New Zealand during 2020 (95% credible interval: 0.243 – 16.0%) but depending on assumptions 39.0% or 37.7% of Ebola infections were not detected in Guinea (95% credible intervals: 1.69 – 87.0% or 1.7 – 80.9%).

## INTRODUCTION

There are many non-pharmaceutical interventions for controlling infectious disease epidemics. Some control measures, such as case isolation and safe and dignified burials avoid secondary infections but others, such as contact tracing, avoid tertiary infections. Measures, which avoid secondary infections, are most effective when tertiary infections are also avoided and all (or nearly all) infections are identified so that interventions can be targeted (1). If contact tracing is implemented well, contacts of known cases can take precautions to reduce onward transmission by limiting their contacts and isolating quickly on symptom onset (2–4). However, if many infections are not detected, outbreaks can grow rapidly as undetected infections usually infect more people than detected cases (5).

Infections or deaths may not be reported for a variety of reasons (6). Poor availability of tests at the start of an outbreak of an emerging pathogen, such as SARS-CoV-2, may mean that those with symptoms cannot be diagnosed (7). Asymptomatic individuals may also not know they are infected unless tested for other reasons, such as through contact tracing (8). Undetected infections are not unique to SARS-CoV-2 and under-reporting is common in Ebola outbreaks due to barriers to accessing health care and limited hospital capacity (9). Many patients may not seek health care due to mistrust and if they die, may be buried without notification, leading again to those cases being missed from official lists (10).

Infectious disease analysis and modelling are important tools for managing epidemics and can help provide quantitative evidence and situational awareness to public health responses (11). The importance of such analyses has been highlighted by the response to the COVID-19 pandemic, which has been, to a large extent, informed by epidemic modelling e.g. (12– 14). However, these models often require robust case data to make accurate transmission predictions. Over time attempts have been made to account for under-reporting in models. Some models assume perfect reporting (15,16), however, this can lead to an underestimation of the infection rate (6). Other methods assume a constant under-reporting rate (17), use data augmentation techniques (6) or rely on more complex models to merge multiple data streams through evidence synthesis (18). More recently, many models have switched to using death data, which was believed to be more reliable than case data, because it is more likely consistent over time and between countries (13). This is especially important for methods which are robust to constant under-reporting.

We propose using a quasi-Bayesian next generation matrix (NGM) approach in this paper to estimate the proportion of infections that are not detected in an outbreak. This method is not disease specific, is simple to implement from contact tracing and surveillance data and can be repeated throughout the outbreak to provide time varying estimates. We investigate the suitability of our method using simulated data and present two applications of our method: the SARS-CoV-2 outbreak in New Zealand (NZ) in 2020 and the Ebola epidemic in Guinea in 2014.

## METHODS

NGMs are often used to calculate the basic reproduction number (the average number of secondary infections generated by a primary infection in a large fully susceptible population), *R*_0_, from a finite number of discrete categories that are based on epidemiologically relevant traits in the population, such as infected individuals at different stages of infection (e.g. exposed and infectious) or with different characteristics (e.g. age) e.g. Baguelin et al. (19). The NGM is a matrix which quantifies the number of secondary infections generated in each category by an infected individual in a given category. *R*_0_ is defined as the dominant eigenvalue of this matrix (20,21). They have also been used by Grantz et al. (22) to evaluate contact tracing systems. Similarly, here we stratify infected individuals using information about their contact tracing status and whether they were being followed up at the time of symptom onset to assign infection pathways and construct our NGM. We identify three types of infections: i) infections that are not detected (ND), ii) infections (or cases) that are detected but not under active surveillance (NAS), and (iii) infections (or cases) that are detected and under active surveillance (AS).

Contact follow-up or surveillance might take different forms for different diseases; for Ebola, a contact under active surveillance would be undergoing in-person follow-up for 21 days after their last interaction with the case (23), whereas for SARS-CoV-2 in some settings, a contact under active surveillance may be notified by contact tracers, or through a mobile phone application, and asked to self-isolate for up to 10 days (24,25).

### Formulation of the NGM

For contact tracing to be fully effective, the parent (or primary) case needs to be diagnosed and, if positive, all their contacts placed under active surveillance. The parent case therefore needs to know and remember everyone they have been in close contact with whilst they have been infectious and for these contacts to be contacted. Despite a contact being recalled and reported, they may not be under active surveillance if they cannot be identified due to missing or incorrect contact details or evasion from contact tracers. We assume in our model that: i) infections that are not detected and those cases detected but not under active surveillance have the same effective reproduction number (*R*) and therefore on average, infect the same number of secondary cases; and ii) AS have a lower effective reproduction number (scaled by *α*) because they are rapidly isolated after the onset of symptoms. We define *ϕ* as the proportion of contacts recalled, *γ* as the proportion of contacts actively under surveillance, and *π* as the proportion of cases detected or “re-captured” by community surveillance.

We identify 12 pathways through which individuals can become infected (Figure 1). These pathways are described as follows:

**Figure 1:**
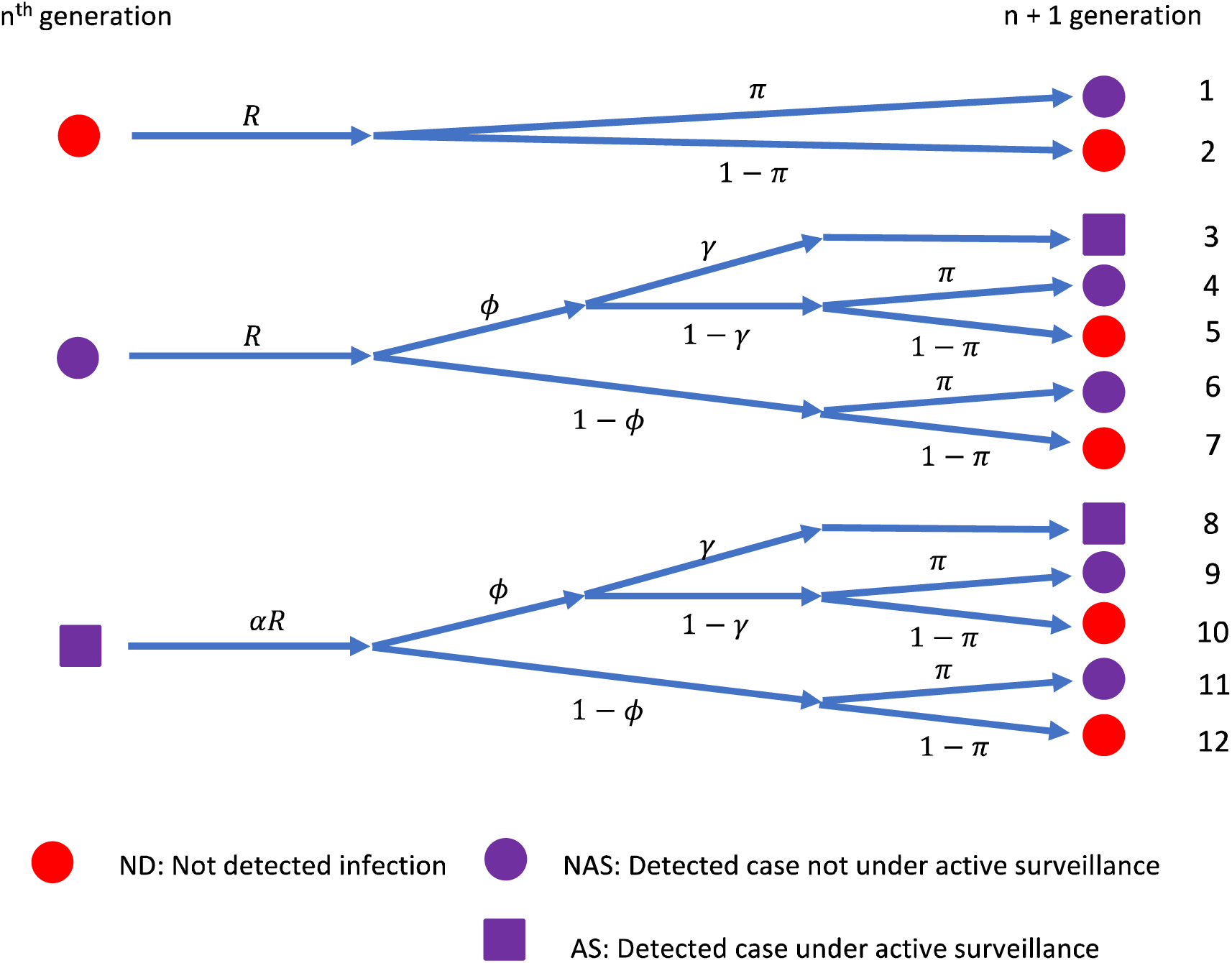
Potential pathways for a three-state model of Ebola surveillance (ND, AS, NAS). R is the effective reproduction number, α is the scaling of the reproduction number due to active surveillance (rapid isolation upon symptom onset), ϕ is the proportion of contacts recalled and reported by a case, γ is the proportion of contacts actively under surveillance, and π is the proportion of cases detected or “re-captured” by community surveillance. We assume that all cases under active surveillance are detected. The coloring and shape of the end points of the paths are described as follows: red circle - any case that was not detected (so cannot be under active surveillance), purple circle - an eventually detected case that was not under active surveillance at the time of symptom onset (e.g. a contact of an earlier case lost to follow-up or who refused follow-up), purple square: a detected case that was under active surveillance at the time of symptom onset (e.g. a contact of a previously detected case, correctly recalled and reported, and under surveillance).

1. A case that was detected (with probability *π*), who was infected by an infection that was not detected and was therefore not under active surveillance.
2. An infection that was not detected (with probability 1-*π*), who was infected by an infection that was not detected and was therefore not under active surveillance.
3. A case that was detected (with probability *π*), who was infected by a case that was detected but not under surveillance, was correctly recalled as a contact (with probability *ϕ*) and was under active surveillance (with probability *γ*).
4. A case that was detected (with probability *π*), who was infected by a case that was detected but that was not under surveillance, was correctly recalled as a contact (with probability *ϕ*) but was not under surveillance (with probability 1-*γ*).
5. An infection that was not detected (with probability 1-*π*), who was infected by a case that was detected but not under surveillance, was correctly recalled (with probability *ϕ*) but was not under surveillance (with probability 1-*γ*).
6. A case that was detected (with probability *π*) case, who was infected by a case that was detected but not under surveillance, that was not recalled (probability 1-*ϕ*).
7. An infection that was not detected (with probability 1-*π*) case, who was infected by a case that was detected but not under surveillance, that was not recalled (probability 1-*ϕ*).
8. A case that was detected (with probability *π*), who was infected by a case that was detected and under surveillance, was correctly recalled (with probability *ϕ*) and was under surveillance (with probability *γ*).
9. A case that was detected (with probability *π*) case, who was infected by a case that was detected and under surveillance, was correctly recalled (with probability *ϕ*) but was not under surveillance (with probability 1-*γ*).
10. An infection that was not detected (with probability 1-*π*), who was infected by a case that was detected and under surveillance, was correctly recalled (with probability *ϕ*) but was not under surveillance (with probability 1-*γ*).
11. A case that was detected (with probability *π*), who was infected by a case that was detected and under surveillance, that was not recalled (with probability 1-*ϕ*).
12. An infection that was not detected (with probability 1-*π*) case, who was infected by a case that was detected and under surveillance, that was not recalled (with probability 1-*ϕ*).

Seven of our twelve pathways result in detected cases. The cases from pathways 3, 4, 8, and 9 are individuals on contact lists who are detected as cases whereas, the cases from pathways 1, 6, and 11 are de novo cases that are not on any contact tracing list, but which are detected via other routes such as attending a health care unit. The cases from pathways 3 and 8 are contacts who were under surveillance at the time of symptom onset, while those from pathways 4 and 9 were not under surveillance at onset. The infections resulting from the pathways 2, 5, 7, 10 and 12 are not detected by the surveillance system. We use the notation F_X_ to denote the likelihood of a case stemming from pathway X, for example F_1_ equals *Rπ*

If *Z*_*n*_ = [*ND*_*n*_, *NAS*_*n*_, *AS*_*n*_]^*T*^ is a vector of the number of each type of case for generation *n*, the dynamics of the model is given by:

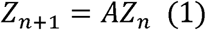

where *A* is our NGM that represent the potential transitions from one generation of cases to the next

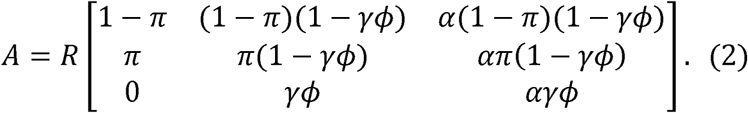

From the eigenvalues of this NGM, we can calculate the proportion of each of the three types of infections (*ND, NAS* and *AS*), see Supplementary Information (SI) A. In the limit as *n* goes to infinity, an equilibrium is reached and the proportion of cases that are not detected, *μ*_*ND*_, can be calculated as:

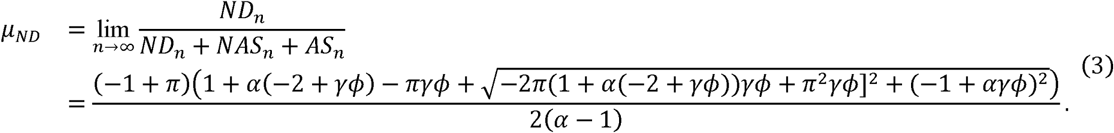

As shown in the calculation in the SI and illustrated Figure S1 in SIA, convergence to this equilibrium value is fast.

### Linking our model to contact tracing and surveillance system data

Cases are often recorded in line-lists during disease outbreaks, where dates of testing, symptom onset and hospitalization are recorded alongside information about the age and sex of the patient. When case lists are linked to contact lists, we can derive two ratios with which we parameterize our NGM. We define *r*_1_ as the ratio of cases who were contacts but not under surveillance versus the cases who were contacts and under surveillance and *r*_2_ as the ratio of de novo cases (cases that were not known contacts) versus detected cases that were contacts and under surveillance.

Following the pathways in Figure 1, we expand *r*_1_ (the ratio of cases who were contacts but not under surveillance versus the cases who were contacts and under surveillance) as 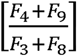. At the equilibrium of the surveillance process (SIA), we have *ND*_*n*_ = *μ*_*ND*_ *C*_*n*_, *NAS*_*n*_ = *μ*_*NAS*_ *C*_*n*_ and *AS*_*n*_ = *μ*_*AS*_ *C*_*n*_, where *C*_*n*_ = *ND*_*n*_ + *NAS*_*n*_ + *AS*_*n*_ is the total number of cases at generation *n, μ*_*NAS*_ is the proportion of cases not under active surveillance and *μ*_*AS*_ is the proportion of cases under active surveillance. Therefore,

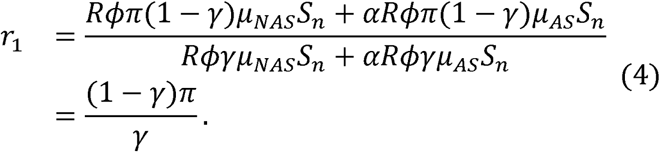

We re-write this as

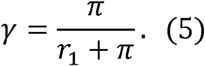

We also expand *r*_2_ (the ratio of de novo cases versus detected cases that were contacts and under surveillance) as 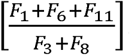. Therefore,

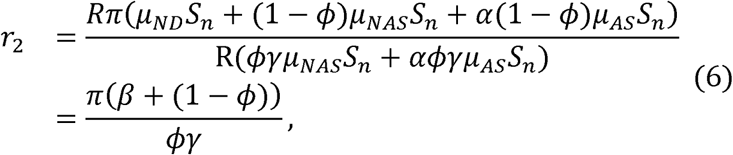

where 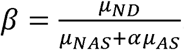. This can be rewritten as

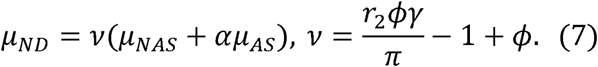

Figure 2 illustrates the dependencies between these two ratios and the parameters in our model in a directed acyclic graph where the green nodes are our data, blue nodes are model parameters and white nodes are calculated parameters.

**Figure 2:**
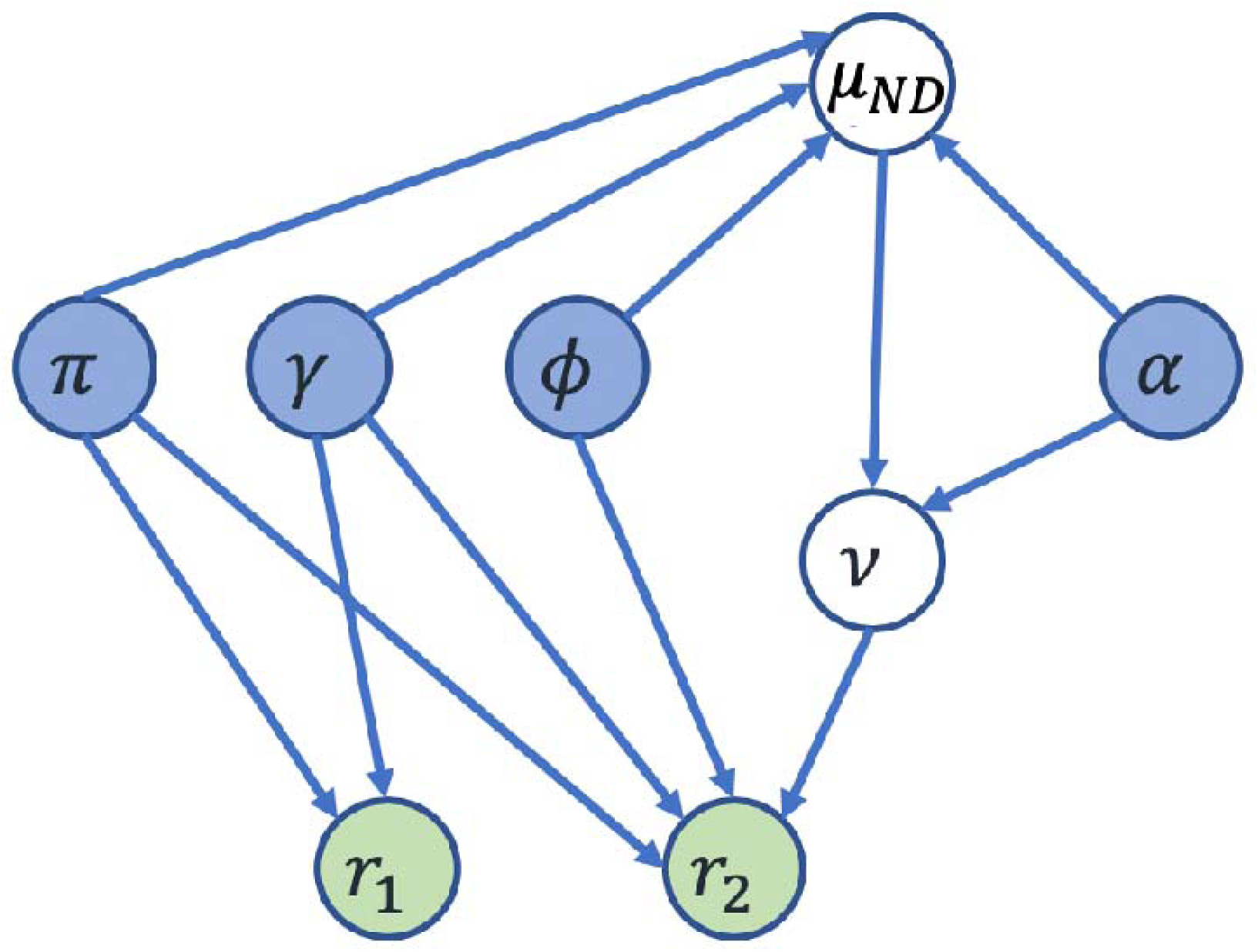
Directed acyclic graph showing the functional relationships of the surveillance model and the ratios observed in the surveillance. The blue nodes represent the parameters of the model that we want to infer (is the proportion of cases detected or “re-captured” by community surveillance; is the proportion of contacts actively under surveillance; is the proportion of contacts recalled by a case and is the scaling of reproduction number due to active surveillance (rapid isolation upon symptom onset)). The green terminal nodes are the potentially observable data (is the ratio of cases who were contacts but not under surveillance versus the cases who were contacts and under surveillance; and as the ratio of de novo cases versus detected cases that were contacts and under surveillance. The white nodes are our calculated terms (is the proportion of cases that are not detected; and relates the proportion of not detected cases to the other two types of cases). The arrows show the direction of the dependence.

In addition to equations (5) and (7), we also have three more relationships that we can use: the proportions of each type of case (*μ*_*ND*_, *μ*_*NAS*_ and *μ*_*AS*_) that are found using the leading eigenvector of the NGM (see SIA). We therefore have five equations and seven unknown parameters (*π, α, ϕ, γ, μ*_*ND*_, *μ*_*NAS*_, *μ*_*AS*_). If we fix two parameters, we can then estimate the other parameters. We choose here to fix *α* since this could be estimated from additional data such as serology and *π*.

### Application to the estimation of the proportion of infections that were not detected

We estimated the proportion of infections that are not detected using a quasi-Bayesian framework for each scenario. For each run of each scenario, we sampled 10,000 values from [0,1]^2^ uniformly for (*π, α*), which is comparable to assuming a uniform prior distribution, and computed the other parameters (*γ, ϕ, μ*_*ND*_)if a solution was viable. We note that there is no solution for some values of (*π, α*)(see SIB). Our credible intervals (CrI) reflect the values between which 95% of our viable samples lie.

#### Simulated data

We investigate the suitability of our method using an individual-based model developed using NetLogo(26) (see SIC) for 3 scenarios:

1. Contact tracing similar to SARS-CoV-2 example in New Zealand (NZ);
2. Contract tracing similar to Ebola in Guinea;
3. Contact tracing similar to Ebola in Guinea and then improves to match the SARS CoV-2 example in NZ after 500 days.

For each scenario, we simulated 1000 runs and sampled each run 10,000 times. Here we assumed prior knowledge about the values of *π* and *α* so uniformly sampled between 0.2 above and below the true values of *π* and *α* (see SIC for parameter value). We compared the probability that the true parameters in each of our scenarios lie within the 95% CrI estimates. We consider two time periods for scenario 3, before and after the parameter change.

We also undertook a sensitivity analysis to investigate relaxing our assumption on *α*, where we compared the estimated values of missing cases when we varied the reduction in the scaling for a NAS case. We compared the probability that the true value of the proportion of infections that were not detected lies within our 95% CrI for scenario one with values of alpha for NAS cases of 0.6 and 0.8 and 1.0 (initial scenario one). We again ran 1000 simulations of each and assumed the parameter were equal to the SARS-CoV-2 scenario.

#### SARS-CoV-2 in New Zealand 2020

Well performing contact tracing systems have been partially credited for the success of NZ’s response to the SARS-CoV-2 epidemic in 2020 (27–29). NZ’s Ministry of Health reported 570 locally acquired cases up until 14^th^ December 2020 that had an epidemiological link to a previous case and 90 cases without an epidemiological link (30). We assume that 80% of contacts were under active surveillance before diagnosis, since 80% was determined as the minimum requirement for the NZ system (25). Therefore, we estimate 456 cases were under active surveillance and 114 cases were not. This makes *r*_1_ = 0.25 and *r*_2_ = 0.20.

#### Ebola in Guinea 2014

We use data from Dixon et al. (31), which present contact tracing outcomes from two prefectures in Guinea between the 20th September and 31st December 2014. The authors found that only 45 cases out of 152 were registered as contacts of known cases across Kindia and Faranah prefectures.

Since there is little published data, we consider two scenarios based on different assumptions about *r*_1_ (ratio of contacts not under active surveillance versus contacts under active surveillance).

1. We assume *r*_1_ is equal to 0.2 (five times as many contacts under active surveillance than not under active surveillance, or 5 out of 6 contacts are under active surveillance). This is based on data from Liberia in 2014 and 2015 where, during the same epidemic as Guinea, 27936 contacts were not under active surveillance, whereas 167419 were (32). Since we know the total number of cases on the contact tracing list, 45, and assume *r*_1_ = 0.2, we estimate the number of contacts under active surveillance to be 38 (denominator of *r*_2_). The number of people not on the contact list for the two regions was 107 (numerator of *r*_2_). Therefore, *r*_2_ is equal to 2.85.
2. We assume *r*_1_ is equal to 0.5 (twice as many contacts under active surveillance than not under active surveillance or two thirds of contacts are under active surveillance) to illustrate the impact of a slightly better surveillance system. Since we know the total number of cases on the contact tracing list, 45, and assume *r*_1_ = 0.5, we estimate the number of contacts under active surveillance to be 30 (denominator of *r*_2_). Therefore, *r*_2_ is equal to 3.57.

We again estimated the proportion of infections that are not detected using our quasi-Bayesian framework for both case studies and took 100,000 samples for each case study, sampling *π* and *α* between 0 and 1. All code necessary to implement the analysis is included open source in the “*MissingCases*” R package on GitHub (33).

## RESULTS

### Simulated data

We find that in our three scenarios, the true proportion of infections that are not detected always lie within the uncertainty intervals of the NGM estimates even in scenario 3 where our parameters are not constant. We note this method performs best early in the outbreak when the number of susceptible are large and not in the tail end of the outbreak. However, not all parameters perform consistently well as shown in Table S2, where *γ* only lies within the interval 75.4% of the time in scenario 1 and *ϕ* only 24.6% of the time in scenario 2. We found that performance remained similar if we reduced alpha for NAS cases (Table S3).

### SARS-CoV-2 in New Zealand 2020

We estimate that only 5.26% (95% CrI: 0.245 - 16.0%) of cases were not detected during this wave of the SARS-CoV-2 pandemic in NZ (see Table 1 for all parameter estimates), which suggests a well-functioning and rigorous contact tracing and surveillance system in NZ. In Figure 3, we find that this estimate comes from a feasible parameter space that is focused along the right-hand side of the parameter space, where the proportion of cases detected in the community (*π*) is high. However, we do not learn anything about the scaling in transmission for traced cases so the uncertainty intervals in the proportion of not detected infections account for this.

**Table 1:** Estimates of the parameters for SARS-CoV-2 in New Zealand

| Parameter | Description | Median estimates (95% CrI) |
| --- | --- | --- |
|  | Proportion of cases detected in the community | 84.8% (61.9, 99.2) |
|  | Scaling of the reproduction number for traced cases | 50.2% (4.28, 98.3) |
|  | Proportion of contacts recalled | 91.9% (86.6, 99.5) |
|  | Proportion of contacts under active surveillance | 77.2% (71.2, 79.9) |
|  | Proportion of infections not detected | 5.26% (0.243, 16.0) |

**Figure 3:**
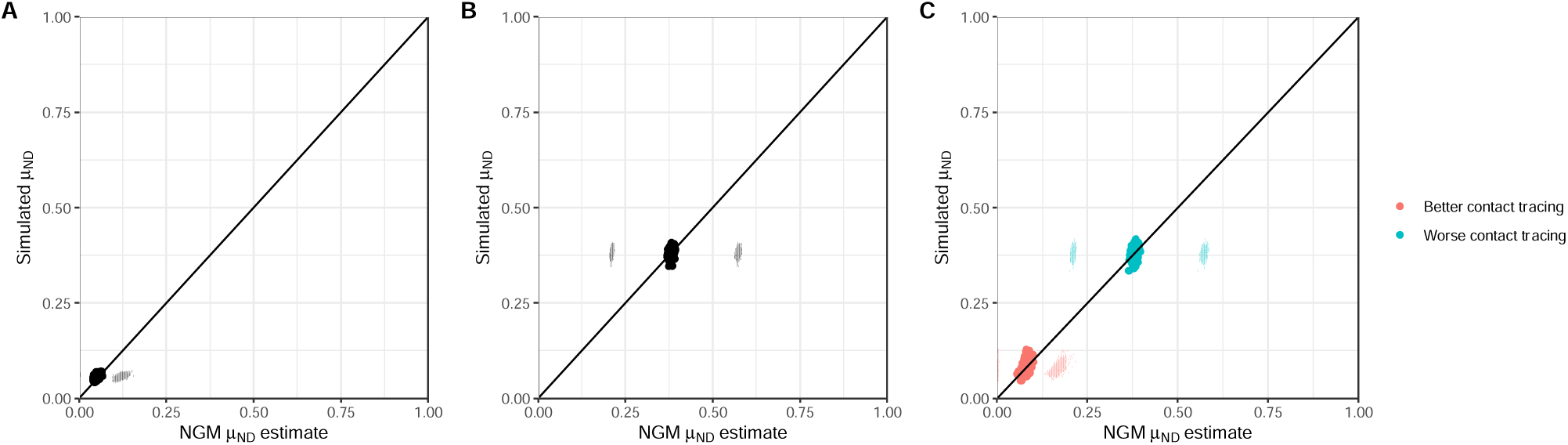
Comparison of NGM estimate of proportion of infections not detected against simulated proportion for 3 scenarios. The error bars parallel to the x-axis depict the 95% CrIs from the NGM estimates. Figure 3A shows a scenario with contact tracing like SARS-CoV-2 in NZ, Figure 3B shows a scenario with contact tracing like Ebola from Guinea and Figure 3C shows a scenario in which contact tracing starts like the Ebola scenario and improves to be like the SARS-CoV-2 scenario. The colors in Figure 3C refer to the two different time periods considered (worse contact tracing: days 100 to 500, better contact tracing days 500 to 900) in our scenarios.

**Figure 4:**
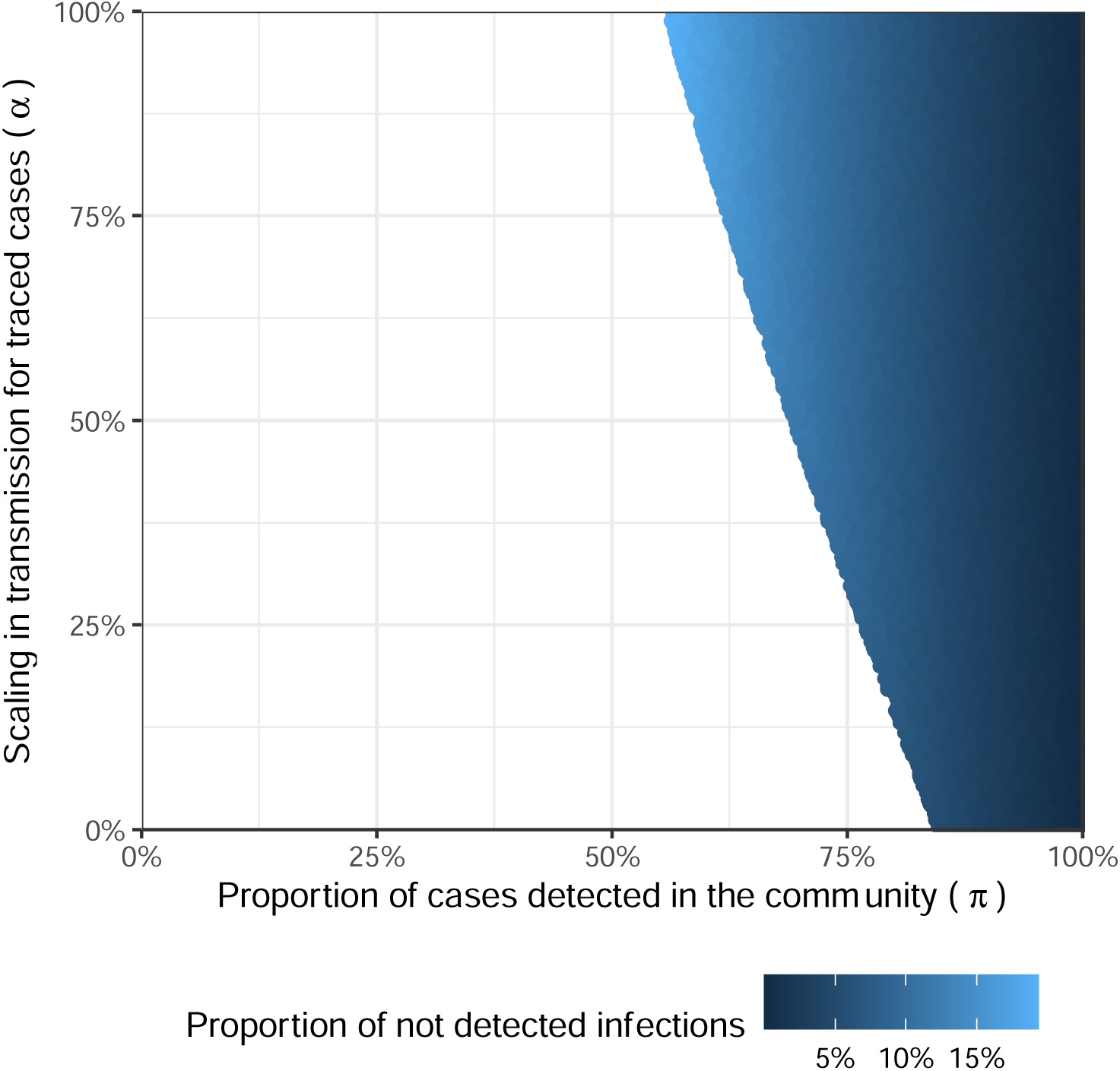
Region of the parameter space compatible with the observed data New Zealand. Values of *π* and *α* are sampled uniformly from [0,1]^2^. The dots show our feasible samples with the color indicating the proportion of not detected infections.

**Figure 5:**
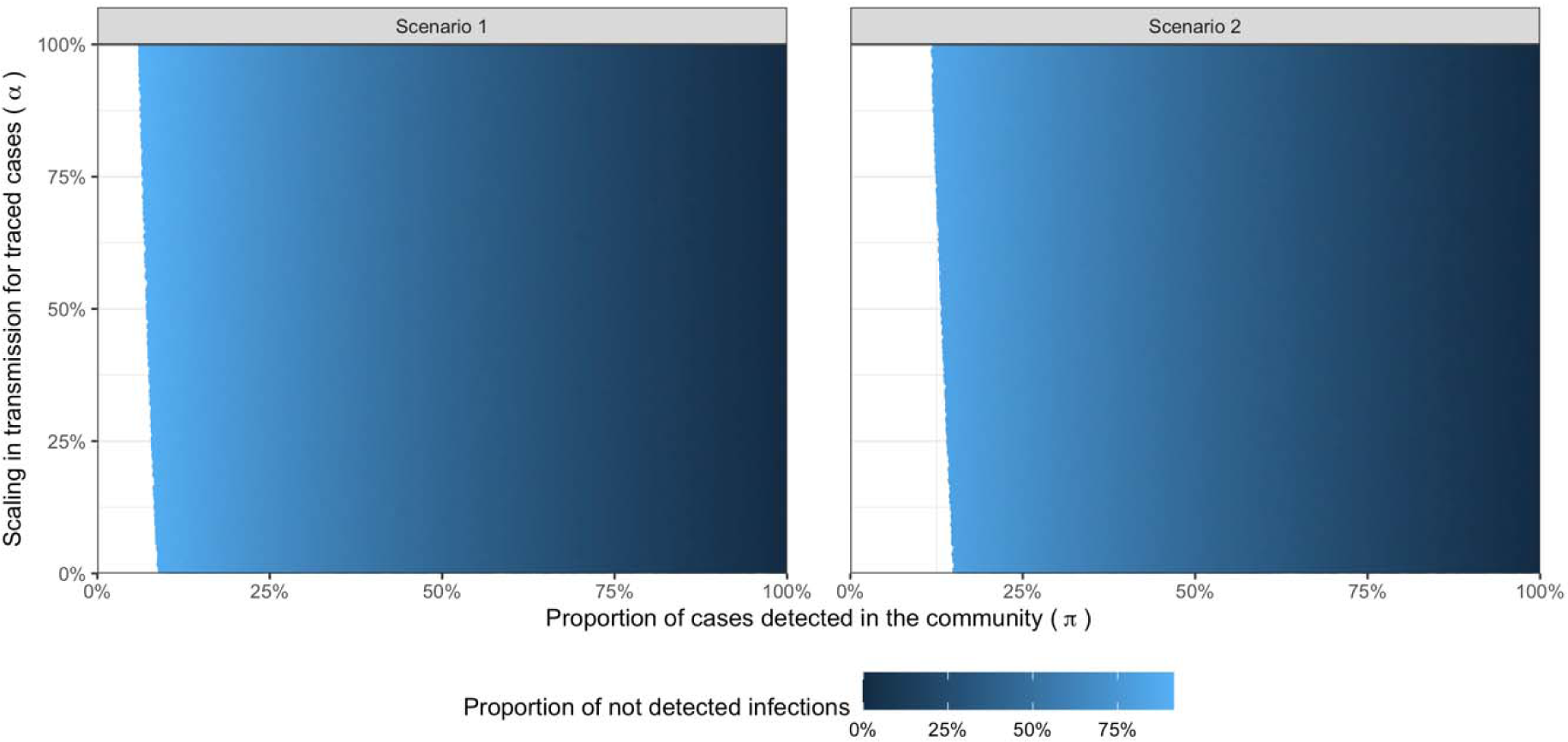
Region of the parameter space compatible with the observed data for the two scenarios in Guinea. Values of and are sampled uniformly from. The dots show our feasible samples with the color indicating the proportion of not detected infections.

### Ebola in Guinea 2014

We estimate that the proportion of Ebola cases that were not detected in Guinea was 39.0% (95% CrI: 1.69-87.0%) or 37.7% (95% CrI 1.70 – 80.9%) for our two scenarios where *r*_1_ = 0.2 and *r*_1_ = 0.5 respectively. The corresponding model parameter estimates for both scenarios are given in Table 2. The only parameter that differs substantially between our scenario is the proportion of contacts under active surveillance, which is directly impacted by the ratio of contacts not under active surveillance versus contacts under active surveillance. We find that we do not learn much about the feasible values of *α* and *π* for these scenarios but as proportions of cases detected in the community fall, the proportion of not detected infections increases.

**Table 2:** Estimates of the parameters for Ebola in Guinea

| Parameter | Description | Median estimates (95% CrI) |  |
| --- | --- | --- | --- |
| | | Scenario 1 ($r_1 = 0.2$) | Scenario 2 ($r_1 = 0.5$) |
| $r_2$ | Ratio of de novo cases versus detected cases that were contacts and under surveillance | 2.85 | 3.57 |
| $\pi$ | Proportion of cases detected in the community | 54.0% (10.1, 97.8) | 57.03% (16.0, 97.9) |
| $\alpha$ | Scaling of the reproduction number for traced cases | 49.8% (2.51, 97.4) | 49.7% (2.54, 97.6) |
| $\phi$ | Proportion of contacts recalled | 35.7% (29.8, 83.1) | 38.6% (29.9, 89.1) |
| $\gamma$ | Proportion of contacts under active surveillance | 73.0% (33.6, 83.0) | 53.3% (24.2, 66.2) |
| $\mu_{ND}$ | Proportion of infections not detected | 39.0% (1.69, 87.0) | 37.7% (1.70, 80.9) |

## DISCUSSION

Contact tracing is an important control mechanism for infectious disease outbreaks. However, its efficiency depends on detecting as many cases as possible. We show in this paper that NGMs can be easily used to estimate the proportion of cases that were not detected in simulated examples and two different disease outbreaks. Our method requires much less data to parameterize our model that other methods, such as capture re-capture (10), which is an alternative method suggested for estimating under-reporting and is highly data intensive. This means that it is feasible to repeat this analysis in near real time as the epidemic unfolds. We find that in our time varying simulation (scenario 3) 95.4% of the simulated proportion of infections not detected lie within the 95% credible intervals but there is a slight bias in the “transient phase” (group 3) where the NGM estimates are higher than the true estimates. This could be because equilibrium had not been reached.

During the West African Ebola epidemic, the WHO acknowledged that their reported case and death figures “vastly underestimate(d)” the true magnitude of the epidemic (34). We find that our estimates for the proportion of cases not detected in Guinea (39.0% (95% CrI: 1.69-87.0%) or 37.7% (95% CrI 1.7 – 80.9%) for our two scenarios where *r*_1_ = 0.2 and *r*_1_ = 0.5 respectively) are in line with values in the literature for neighbouring countries. Dalziel et al. (9) suggested reporting rates in Sierra Leone of 68% (32% under reporting) in the Western Area Urban on 20 October 2014 using burial data. However, higher under reporting has also been estimated: the US Centers for Disease Control and Prevention (35) estimated a 40% reporting rate (60% under-reporting) from Ebola treatment unit bed data and Gignoux et al. (36) estimated a 33% (67% under-reporting) from a capture and recapture study in Liberia between June and August 2014.

Our estimates of the proportion of cases that were not detected during the SARS-Cov-2 outbreak in NZ of 5.26% (95% CrI 0.243 - 16.0%) is in-line with the good health care facilities and the low community transmission of SARS-CoV-2 in NZ (30), but we did not find any estimates in literature to compare our estimates to.

A benefit of this method is that we do not just estimate the proportion of cases that were not detected but also other useful quantities that are important for managing a response such as the proportion of contacts recalled and under surveillance. The wide CrI, especially in second and third simulated data scenarios and the Ebola case study, come from the uniform sample of (*π, α*). This is a limitation of the method but could be improved with better understanding of the performance of the routine surveillance (*π*) and changes in transmissibility due to contact tracing status (*α*) which would narrow the region in the parameter space. A second limitation is our assumption on *α* that only detected cases under active surveillance have a reduced transmissibility. In this simple framework, it is not possible to relax this assumption; however if additional information such as serology was available, we believe this could be used to form a prior distribution on this parameter and potentially allow users to further vary the number of people NAS and ND individuals infect or improve the accuracy of some of the other parameter estimates. As we see in our sensitivity analysis, this does not impact our estimation of the proportion of infections that were not detected but potentially other parameters. A third limitation is that we do not account for differing times to locate contacts within each group, which would further vary the number of cases each case goes on to infect.

We believe this method highlights important lessons for responding to the ongoing SARS-CoV-2 pandemic and the unfortunate inevitability of future infectious disease outbreaks. By simply linking the case line-lists and contact tracing lists, we can use the very general method from our “MissingCases” package (33) to assess under-reporting throughout an epidemic. This would help outbreak responses, especially during the early and late phases, target resources and quantify how effect their surveillance systems were. In addition, these estimates can be used to improve the accuracy of other models, such as for the time varying reproduction number, which are key tools for the outbreak response themselves.

## Data Availability

The data used is included in the paper and code can be found at https://github.com/mrc-ide/MissingCases.

https://github.com/mrc-ide/MissingCases

## Acknowledgments

All authors acknowledge funding from the MRC Centre for Global Infectious Disease Analysis (reference MR/R015600/1), jointly funded by the UK Medical Research Council (MRC) and the UK Foreign, Commonwealth & Development Office (FCDO), under the MRC/FCDO Concordat agreement and is also part of the EDCTP2 programme supported by the European Union; and acknowledge funding by Community Jameel. HJTU also acknowledges funding from Imperial College, London for her fellowship. CAD also acknowledge the NIHR Health Protection Research Unit in Emerging and Zoonotic Infections.

