## Supplementary Information for "Using next generation matrices to estimate the proportion of infections that are not detected in an outbreak"

### Supplementary Information A: Convergence of the surveillance system

Here we derive the proportion infections that are not detected using a framework where the proportion of cases that are not detected does not directly depend on the values of the reproduction number, $R$. Following on from equation 1, we re-write

$$Z_{n}=\prod_{i=0}^{n} RBZ_{0}, (S1)$$

where if we assume $\xi=\gamma\phi$,

$$B=\left[ \begin{matrix} 1-\pi& (1-\pi)(1-\xi) & \alpha(1-\pi)(1-\xi) \\ \pi& \pi(1-\xi) & \alpha\pi\left( 1-\xi\right) \\ 0 & \xi& \alpha\xi\end{matrix} \right]. (S2)$$

Assuming $B$ is diagonalisable, we can rewrite equation S2 as:

$$B=\lambda_{1}P\left[ \begin{matrix} 1 & 0 & 0 \\ 0 & \frac{\lambda_{2}}{\lambda_{1}} & 0 \\ 0 & 0 & \frac{\lambda_{3}}{\lambda_{1}} \end{matrix} \right]P^{-1} (S3)$$

where $\lambda$denote eigenvalues of which $\lambda_{1}$ is the biggest, and

$$P=\left[ \begin{matrix} a & d & g \\ b & e & h \\ c & f & i \end{matrix} \right], (S4)$$

is the matrix composed of the eigenvectors $[a,b,c]^{⊺}$, $[d,e,f]^{⊺}$ and $[g,h,i]^{⊺}$ in order of descending eigenvalues. Following this decomposition, the evolution of the system can be rewritten as:

$$Z_{n}=\lambda_{1}^{n}\prod_{i=0}^{n} RP\left[ \begin{matrix} 1 & 0 & 0 \\ 0 & \left( \frac{\lambda_{2}}{\lambda_{1}} \right)^{n} & 0 \\ 0 & 0 & \left( \frac{\lambda_{3}}{\lambda_{1}} \right)^{n} \end{matrix} \right]P^{-1}Z_{0}. (S5)$$

We define

$$\left[ \begin{matrix} j \\ k \\ l \end{matrix} \right]=P^{-1}Z_{0} (S6)$$

as the vector of the initial conditions in the coordinate system defined by the $P^{-1}$ matrix.

Therefore, we can re-write equation S5 as

$$\begin{matrix} Z_{n} & =\lambda_{1}^{n}\prod_{i=0}^{n} RP\left[ \begin{matrix} 1 & 0 & 0 \\ 0 & \left( \frac{\lambda_{2}}{\lambda_{1}} \right)^{n} & 0 \\ 0 & 0 & \left( \frac{\lambda_{3}}{\lambda_{1}} \right)^{n} \end{matrix} \right]\left[ \begin{matrix} j \\ k \\ l \end{matrix} \right], \\ & =\lambda_{1}^{n}\prod_{i=0}^{n} RP\left[ \begin{matrix} j \\ \left( \frac{\lambda_{2}}{\lambda_{1}} \right)^{n}k \\ \left( \frac{\lambda_{3}}{\lambda_{1}} \right)^{n}l \end{matrix} \right]. \end{matrix} (S7)$$

This means we can define the number of each type of infection (*ND, NAS* and *AS*) in generation $n$ as

$$\begin{matrix} {ND}_{n} & =\left\{ aj+dk\left( \frac{\lambda_{2}}{\lambda_{1}} \right)^{n}+gl\left( \frac{\lambda_{3}}{\lambda_{1}} \right)^{n} \right\}\lambda_{1}^{n}\prod_{i=0}^{n} R, \\ {NAS}_{n} & =\left\{ bj+ek\left( \frac{\lambda_{2}}{\lambda_{1}} \right)^{n}+hl\left( \frac{\lambda_{3}}{\lambda_{1}} \right)^{n} \right\}\lambda_{1}^{n}\prod_{i=0}^{n} R, \\ {AS}_{n} & =\left\{ cj+fk\left( \frac{\lambda_{2}}{\lambda_{1}} \right)^{n}+il\left( \frac{\lambda_{3}}{\lambda_{1}} \right)^{n} \right\}\lambda_{1}^{n}\prod_{i=0}^{n} R. \end{matrix}$$

and the proportion of infections that are not detected.

$$\frac{{ND}_{n}}{{ND}_{n}+{NAS}_{n}+{AS}_{n}}=\frac{aj+dk\left( \frac{\lambda_{2}}{\lambda_{1}} \right)^{n}+gl\left( \frac{\lambda_{3}}{\lambda_{1}} \right)^{n}}{j(a+b+c)+k(d+e+f)\left( \frac{\lambda_{2}}{\lambda_{1}} \right)^{n}+l(g+h+i)\left( \frac{\lambda_{3}}{\lambda_{1}} \right)^{n}}. (S8)$$

As $\left( \frac{\lambda_{2}}{\lambda_{1}} \right)^{n}<1$ and $\left( \frac{\lambda_{3}}{\lambda_{1}} \right)^{n}<1$ by construction, we have:

$$\lim_{n\to\infty}\frac{{ND}_{n}}{{ND}_{n}+{NAS}_{n}+{AS}_{n}}=\frac{a}{a+b+c}=\mu_{ND}$$

where $[a,b,c]^{⊺}$ is the eigenvector associated with the biggest eigenvalue of $B$. This eigenvector can be found by calculating the determinant of $B-\lambda I$, were $I$ is the 3x3 identity matrix.

The largest eigenvalue of B is equal to

$$\frac{1}{2}\left( 1-\pi\xi+\alpha\xi+\sqrt{(-1+\pi\xi-\alpha\xi)^{2}-4(\alpha\xi-\pi\alpha\xi)} \right)$$

and corresponds to the eigenvector

$$\left[ \begin{matrix} (-1+\pi)\frac{-1+\pi\xi+\alpha\xi-\sqrt{-2\pi(1+\alpha(-2+\xi))\xi+\pi^{2}\xi^{2}+(-1+\alpha\xi)^{2}})}{2\pi\xi} \\ \frac{1-\pi\xi-\alpha\xi+\sqrt{-2\pi(1+\alpha(-2+\xi))\xi+\pi^{2}\xi^{2}+(-1+\alpha\xi)^{2}}}{2\xi} \\ 1 \end{matrix} \right].$$

Therefore,

$$\mu_{ND}=\frac{(-1+\pi)(1+\alpha(-2+\xi)-\pi\xi+\sqrt{-2\pi(1+\alpha(-2+\xi))\xi+\pi^{2}\xi]^{2}+(-1+\alpha\xi)^{2}})}{2(\alpha-1)}. (S9)$$

We now show an example of the convergence of the system. We assume $\pi=$ 0.848, $\alpha=$ 0.484, $\gamma=$ 0.772 and $\phi=$ 0.919. We chose a distribution for $R$ to get some variability while keeping a relatively contained epidemic.

$$R\sim\mathcal{U}(0.2,2)$$

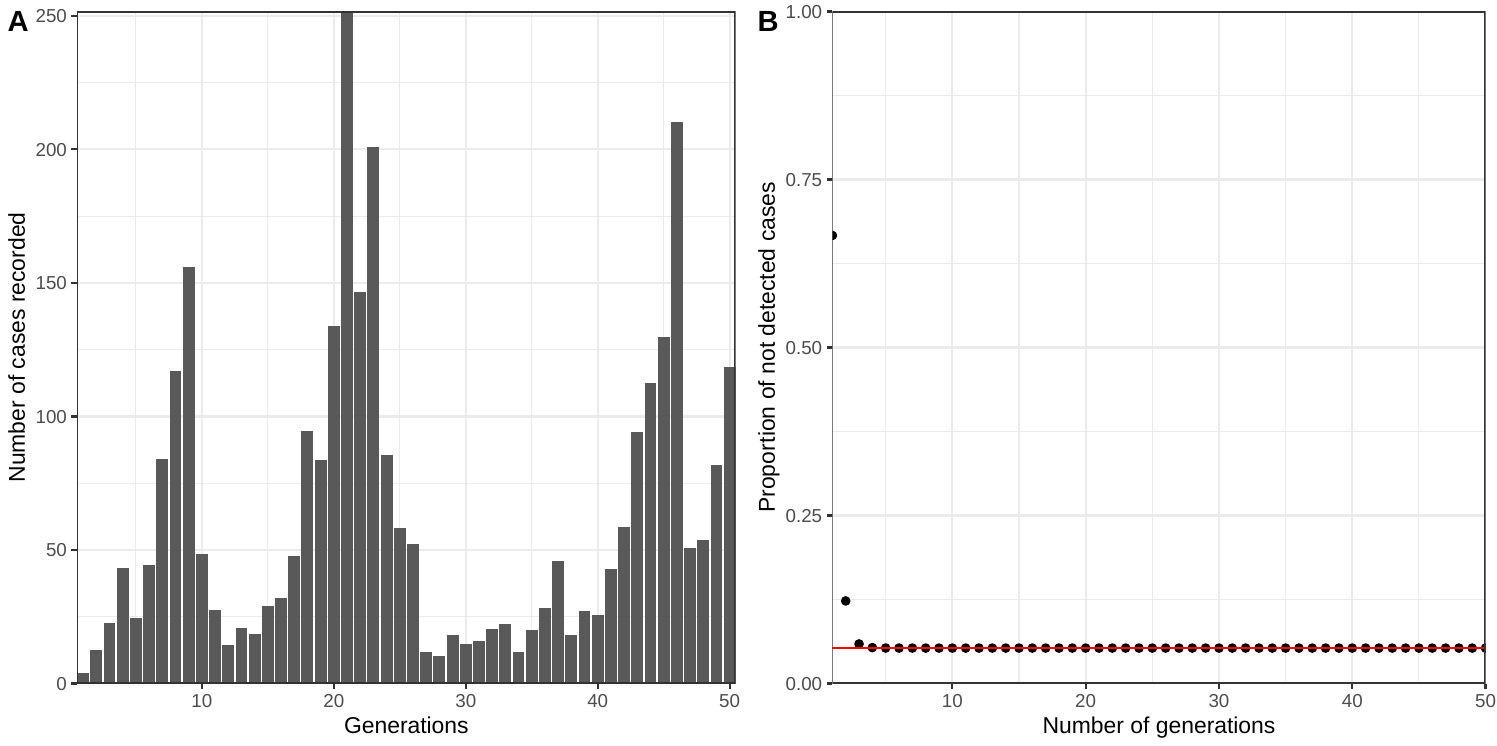

Figure S1: Evolution of simulated outbreak. A) Number of cases for each generation of the example model where $\pi=0.848$, $\alpha=0.484$, $\gamma=0.772$, $\phi=0.919$ and $R\sim\mathcal{U}(0.2,2)$. B) Proportion of not detected infections over a number of generations. The black dots are values derived from simulation while the red line shows the value derived analytically.

We see that, as predicted by the analytic derivation, despite the unpredictable course of the epidemics (Figure S1A), the proportion of infections that are not detected quickly reaches an equilibrium (Figure S1B). This proportion at equilibrium can be calculated using the formula derived above. We note that the convergence is exponential and thus fast proving that $\lambda_{1}\gg\lambda_{2}$ and $\lambda_{1}\gg\lambda_{3}.$

### Supplementary Information B: Solution of the system of equations assuming parameters

We assume that $\pi=$ 0.848, $\phi=$ 0.919 and get $r_{1}=$ 0.25 and $r_{2}=$ 0.20 from the NZ data. Using Equation 5, we estimate $\gamma$ = 0.772. Solving the system gives the following solution: $\alpha=$ 0.484 and $\mu_{1}=$ 0.053 meaning that in this configuration, 5% of the infections are not detected and missing from the records. A graphical representation is given in Figure S2, where the solution of the system can be seen at the intersection of the two curves. No solution is found if the two curves do not intersect.

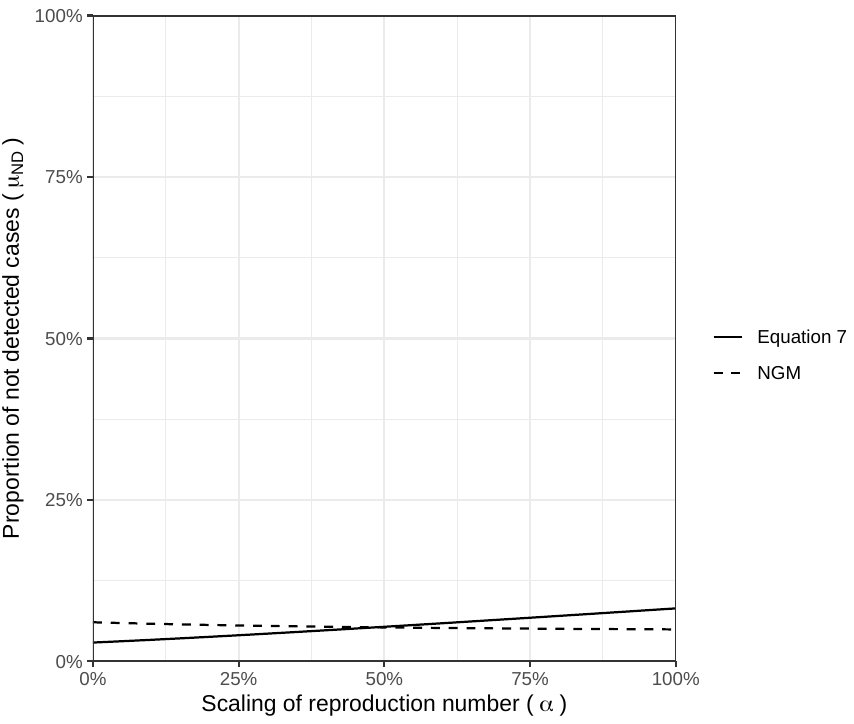

Figure S2: Derivation of $\alpha$ the scaling of the reproduction number following contactt tracing and $\mu_{ND}$ the proportion of not detected cases, assuming $\pi=0.848$ and $\phi=0.919$. The solid line links $\mu$ and $\alpha$ by using the proportion of de novo cases observed vs cases who are contact and followed up. The dashed line indicates the relationship given by the dynamics of the next generation model.

**Supplementary Information C: NetLogo individual-based model**

We use the NetLogo software to develop a simple. individual-based model with susceptible, exposed, infectious and recovered individuals. For all scenarios, we assume it takes 20 days after someone got exposed to become infectious and that duration of infection lasts 80 days. 996 individuals begin each simulation as susceptible, with four split between four exposure classes (1 in each) depending on their contact tracing status:

1) Individuals that were exposed to infection and will never be detected.
2) Individuals that were exposed to infection when they were not on the contacts list and found by routine surveillance.
3) Individuals that were exposed to infection when they were on the contact list but were not under active surveillance and found by routine surveillance.
4) Individuals that were exposed to infection when they were under active surveillance.

We use the diagram in Figure 1 along with assumed values for our parameters to determine the evolution of the outbreak in each scenario up to 900 steps. At this point most outbreaks were over. During each simulation, we tracked the number of individuals in the four categories above so we could calculate the ratios and the proportion of infections that were not detected. We assumed once a case is recovered, it cannot be infected again.

We choose parameters to mimic the examples in the paper:

1) Contact tracing similar to SARS-CoV-2 example in New Zealand (NZ);
2) Contract tracing similar to Ebola in Guinea;
3) Contact tracing similar to Ebola in Guinea and then improves to match the SARS-CoV-2 example in NZ after 500 days.

The values of each parameter in our 3 scenarios and sampling ranges can be found in Table S1 and the NetLogo examples are included in our GitHub (<https://github.com/mrc-ide/MissingCases>).

For scenario 3, we grouped our results into 2 separate periods of 400 days (100 to 499, and 500 to 899) and calculated the number of new infections of each type during each period. We removed the first 100 days of the outbreak to remove the impact of the artificial seeding imposed and the end to ensure elimination had not already been reached in our scenarios. We also removed any time periods where no new infections were generated. For each simulation, we calculated the $r_{1}$ , $r_{2}$ and $\mu_{ND}$for the four periods from the data and ran the NGM method to get each set of parameter estimates. We then saw whether the true parameter estimates lay within the 95% credible intervals.

Table S1: Parameters for our NetLogo scenarios. Sampling ranges for NGM method input arguments given in brackets

| **Parameter** | **Description** | **Scenario 1** | **Scenario 2** | **Scenario 3** |
| --- | --- | --- | --- | --- |
| $\pi$ | Proportion of cases detected in the community | 0.848 (0.648 - 1.00) | 0.540 (0.340 - 0.740) | Scenario 2 parameters until t = 500 then scenario 1 |
| $\alpha$ | Scaling of the reproduction number for traced cases | 0.502 (0.302 – 0.702) | 0.498 (0.298 – 0.698) |  |
| $\phi$ | Proportion of contacts recalled | 0.919 | 0.357 |  |
| $\gamma$ | Proportion of contacts under active surveillance | 0.772 | 0.730 |  |

Table S2: Percentage of scenario evaluations where true parameter lies within 95% credible intervals.

| Parameter | Description | Scenario 1 | Scenario 2 | Scenario 3 |
| --- | --- | --- | --- | --- |
| $\pi$ | Proportion of cases detected in the community | 100.0% | 100.0% | 100.0% |
| $\alpha$ | Scaling of the reproduction number for traced cases | 100.0% | 100.0% | 100.0% |
| $\phi$ | Proportion of contacts recalled | 100.0% | 24,6% | 45.7% |
| $\gamma$ | Proportion of contacts under active surveillance | 75.4% | 99.4% | 87.1% |
| $\mu_{ND}$ | Proportion of cases not detected | 100.0% | 100.0% | 100.0% |

Table S3: Percentage of scenario one evaluations where true parameter lies within 95% credible intervals when $\alpha$ for NAS is 1.0 (model assumption scenario), 0.8 and 0.6.

| Parameter | Description | $\boldsymbol{\alpha}_{\boldsymbol{NAS}}$ = 1.0 | $\boldsymbol{\alpha}_{\boldsymbol{NAS}}$ = 0.8 | $\boldsymbol{\alpha}_{\boldsymbol{NAS}}$ = 0.6 |
| --- | --- | --- | --- | --- |
| $\pi$ | Proportion of cases detected in the community | 100.0% | 100.0% | 100.0% |
| $\alpha$ | Scaling of the reproduction number for traced cases | 100.0% | 100.0% | 100.0% |
| $\phi$ | Proportion of contacts recalled | 100.0% | 100.0% | 100.0% |
| $\gamma$ | Proportion of contacts under active surveillance | 75.4% | 85.8% | 87.8% |
| $\mu_{ND}$ | Proportion of cases not detected | 100.0% | 100.0% | 100.0% |
